# In end-stage kidney disease, inflammation, erythron abnormalities and declined kidney function tests are accompanied by increased affective symptoms, chronic-fatigue, and fibromyalgia

**DOI:** 10.1101/2023.01.12.23284460

**Authors:** Hussein Kadhem Al-Hakeim, Basim Abd Al-Raheem Twaij, Mustafa Hassan Ahmed, Abbas F. Almulla, Shatha Rouf Moustafa, Michael Maes

**Affiliations:** Department of Chemistry, College of Science, University of Kufa, Iraq; Nephrologist at the Nephrology Department, Al-Hakeem General Hospital, Najaf, Iraq; Medical Laboratory Technology Department, College of Medical Technology, The Islamic University, Najaf, Iraq; Clinical Analysis Department, College of Pharmacy, Hawler Medical University, Havalan City, Erbil, Iraq; Department of Psychiatry, Faculty of Medicine, Chulalongkorn University, Bangkok, Thailand; Department of Psychiatry, Medical University of Plovdiv, Plovdiv, Bulgaria; Deakin University, IMPACT, the Institute for Mental and Physical Health and Clinical Translation, School of Medicine, Barwon Health, Geelong, Australia

**Keywords:** mood disoders, major depression, chronic fatigue syndrome, inflammation, oxidative stress, biomarkers

## Abstract

**Background:** Numerous neuropsychiatric symptoms, including affective symptoms, chronic fatigue syndrome, and fibromyalgia symptoms, are present in patients with end-stage renal disease (ESRD). This study examines the relationship between neuropsychiatric symptoms and red blood cell (RBC) parameters, kidney function tests, zinc, C-reactive protein, and calcium levels in patients with ESRD.

**Methods:** The above biomarkers and the Beck-Depression Inventory, the Hamilton Anxiety Rating Scale, and the Fibro-Fatigue Rating Scale were measured in 70 patients with end-stage renal disease (ESRD) and 46 healthy controls.

**Results:** Increased scores of depressive, anxious, cognitive, and physiosomatic symptoms (including chronic fatigue, fibromyalgia, and autonomous symptoms) characterise ESRD. One latent vector could be extracted from these diverse symptom domains, which are, therefore, manifestations of a common core referred to as the physio-affective phenome. The combined effects of aberrations in red blood cells (RBC) (number of RBC, hematocrit, and haemoglobin), kidney function tests (glomerular filtration rate, ureum, creatinine, albumin, and total serum protein), C-reactive protein, zinc, and copper explained 85.0% of the variance in the physio-affective phenome. In addition, the effects of kidney function decline on the phenome were partially mediated by RBC aberrations and elevated copper, whereas the effects of dialysis frequency were entirely mediated by decreased zinc and elevated CRP.

**Conclusions:** Affective (depression and anxiety), cognitive, and physiosomatic symptoms due to ESRD are interrelated manifestations of the physio-affective phenome, which is driven by (in descending order of importance) kidney dysfunctions, erythron deficits, inflammation, elevated copper, and decreased zinc.

## Introduction

End-stage renal disease (ESRD), the late stage of kidney failure when the glomerular filtration rate (GFR) is 15 ml/min/1.73 m^2^ of body surface area or below, requires hemodialysis or kidney transplantation (Abbasi et al., 2010; Levey et al., 2002). As GFR decreases, biological and clinical dysfunctions progress, including disruptions in energetic cellular metabolism, protein malnutrition, changes in nitrogen input/output, insulin resistance, alterations in the erythron, and a significant increase in the synthesis of inflammation/oxidative stress (OS) mediators (Russa et al., 2019). The severity of ESRD depends on decreases in the estimated GFR (eGFR) and the consequent biochemical aberrations including changes in the erythron or red blood cell (RBC) properties and functions, which are affected by the uremic environment (Bonomini et al., 2017).

Anemia, which is accompanied by a shortening of the circulating RBCs’ half-life, is a common complication in patients with ESRD and is associated with poor long-term survival (Coyne et al., 2017). Moreover, hemodialysis can decrease the average survival time of RBCs due to the compression and twisting of the cells (Thorsteinsdottir et al., 2012; Vashistha et al., 2016). Mean cell volume (MCV) is significantly lower in uremic patients than in controls (Buemi et al., 2002), and MCV values higher than 102 fl are associated with increased mortality (Tennankore et al., 2011). Significant increases are observed in corpuscular hemoglobin concentration (MCHC), and corpuscular hemoglobin (MCH) in ESRD patients compared with controls (Gunawickrama et al., 2021). Alterations in MCHC are correlated with the gradual worsening of the eGFR and renal functions (Gunawickrama et al., 2021). In patients undergoing continuous ambulatory peritoneal dialysis (CAPD), higher red cell distribution width (RDW), a parameter that reflects the volumetric heterogeneity of RBCs in peripheral blood, is associated with increased morbidity and mortality (Hsieh et al., 2017).

In addition to the alterations in the RBCs, ESRD is accompanied by many biochemical changes, including in inflammatory mediators, such as C-reactive protein (CRP) (Babaei et al., 2014; Oweis et al., 2021), trace elements including copper and zinc, (Almeida et al., 2020; Dizdar et al., 2020) and OS biomarkers (Sangeetha Lakshmi et al., 2018; Song et al., 2020). Plasma levels of zinc, a strong antioxidant, decline progressively with decreasing GFR, and, in hemodialysis patients, zinc deficiency may reach a prevalence between 40 and 78% (Dvornik et al., 2006; Navarro-Alarcon et al., 2006; Shaikh et al., 2022). Serum copper levels, on the other hand, are increased in hemodialysis patients (Navarro-Alarcon et al., 2006; Shaikh et al., 2022). Malfunctions of electrolyte channels and transporters in the injured kidneys may cause abnormalities in sodium, potassium, chloride, and phosphate (Barbour et al., 2008; Einhorn et al., 2009; Kestenbaum et al., 2005). CKD and ESRD are accompanied by lowered serum calcium levels (Roumeliotis et al., 2020; Timofte et al., 2021).

ESRD patients frequently experience affective symptoms, including depression and anxiety, and physiosomatic symptoms, including fatigue, fibromyalgia, muscular pain, insomnia, headache, and cognitive impairments (Afshar et al., 2012; Aminoff, 2014; Asad et al., 2023; Brown et al., 2017; Hamed, 2019; Karakan et al., 2011; Lee et al., 2007; Yoong et al., 2017). The reported prevalence rate of depression in patients with CKD ranges from 20% to 30% (Cukor et al., 2007; Hedayati and Finkelstein, 2009), with 49.9% of the patients reporting depressive symptoms (Yoong et al., 2017) and 45.4% reporting anxiety symptoms. The prevalence of fatigue in renal disease ranges from 42-89% depending on the rating scales used (Artom et al., 2014).

Compared to healthy controls, depressed patients have significantly lower RBCs, hematocrit (Hct), and haemoglobin (Hb) levels, while RDW and reticulocytes are significantly elevated, indicating inflammation-associated anaemia (Maes et al., 1996; Vandoolaeghe et al., 1999; Wysokinski and Szczepocka, 2018). Compared to RBCs of healthy controls, RBCs of patients with chronic fatigue syndrome (CFS) are less deformable and exhibit lower membrane fluidity and zeta-surface charge (Saha et al., 2019). It is well known that anaemia and iron deficiency can cause symptoms of chronic fatigue.

Lowered serum zinc is a hallmark of depression, severity of depression, and treatment resistance in that illness (Maes et al., 1994; Maes, M. et al., 1997). Lowered serum zinc levels are also detected in patients with CFS (Maes et al., 2006). Reduced serum calcium levels are associated with depressive and CFS-like symptoms which appear in acute COVID-19 infection and Long COVID (Al-Jassas et al., 2022; Maes et al., 2022), whilst increased serum copper is associated with affective symptoms in type-2 diabtes mellitus and CFS-like symptoms in hemodialysis patients (Al-Hakeim et al., 2021; Asad et al., 2023). Nevertheless, there are no data on whether in end-stage ESRD, affective, physiosomatic, fatigue and fibromyalgia symptoms are associated with abnormalities in kidney function tests, disorders in the erythron, lowered zinc and calcium and increased copper levels.

Hence, the present study was conducted to examine whether affective and physiosomatic symptoms in end-stage ESRD may be explained by abnormalities in kidney function tests and the erythron, lowered zinc and calcium levels, and increased copper levels.

## Subjects and Methods

### Subjects

Seventy ESRD patients participated in the current study. All of them had a previous acute kidney injury (AKI), which progressed into the fifth stage of renal failure, and all were on continuous dialysis. The patients were recruited at the Dialysis Unit at Al-Hakeem General Hospital in Najaf Governorate from March 2022 - May 2022. The assessment of patients was carried out based on their full medical history, which considered the presence of any systemic disease. A senior physician diagnosed the patients according to the 10th revision of the International Statistical Classification of Diseases and Related Health Problems (2021 ICD-10-CM Diagnosis Code N18.6). All patients were on continuous treatment with folic acid or iron and folate formula (Fefol^®^) in addition to calcium carbonate, epoetin alfa (Eprex^®^), and heparin. The control group consisted of 46 healthy subjects recruited from the same catchment area; they were sex- and age-matched to the patients. Patients and controls did not show lifetime axis-1 diagnoses of prior neuropsychiatric disorders, including major depressive episodes, anxiety disorders, schizophrenia, bipolar disorder, psycho-organic disorders, substance use disorders (except nicotine dependence), and chronic fatigue syndrome. All participants were also free of medical conditions other than ESRD including diabetes mellitus, inflammatory bowel disease, multiple sclerosis, Parkinson’s or Alzheimer’s disease, stroke, oncologic disorders, and autoimmune disorders.

Written informed consent was obtained from the patients or their first-degree relatives. The institutional ethics board of the University of Kufa (Document No.2229T / 2022) and the Najaf Health Directorate, Training, and Human Development Center (Document No.20123 / 2022) approved the present study. The study was conducted according to Iraqi and international ethics and privacy laws and ethically under the World Medical Association Declaration of Helsinki. Furthermore, our IRB follows the International Guideline for Human Research Protection as required by the Declaration of Helsinki, the Belmont Report, the CIOMS Guideline, and the International Conference on Harmonization in Good Clinical Practice (ICH-GCP).

### Clinical assessments

A senior psychiatrist utilized the FibroFatigue scale (Zachrisson et al., 2002) to rate the severity of fibromyalgia and chronic fatigue syndrome and the Hamilton Anxiety Rating Scale (HAMA) to rate the degree of anxiety (Hamilton, 1959). All participants finished the Beck Depression Inventory (BDI)-II rating scale on the same day (Beck et al., 1996). The same senior psychiatrist used a semi-structured interview to collect sociodemographic and clinical data. The FF, HAMA and BDI-II rating scale items were used to calculate scores for different symptom subdomains. Pure depressive symptoms were the sum of BDI-II items, namely: sadness, discouragement about the future, feeling like a failure, dissatisfaction, feeling guilty, self-disappointment, criticism of oneself, suicidal ideation, crying, loss of interest, difficulty with decisions, looking unattractive, and work inhibition. Pure anxiety was conceptualized as the sum of HAMA items, namely: anxious mood, tension, fears, and anxious behavior at interview. Pure physio-somatic symptoms were conceptualized as a z composite score based on FF, HAMA and BDI-II items, namely: muscle pain, muscle tension, fatigue, autonomic symptoms, gastro-intestinal symptoms, headache, flu-like malaise (all FF), fatigue, loss of libido (BDI-II), somatic muscular, somatic sensory, cardiovascular symptoms, respiratory symptoms, gastro-intestinal symptoms, and autonomic symptoms (all HAMA). The other symptom domains were conceptualized as z unit-based composite scores. Fatigue was the sum of the z scores of fatigue (FF and BDI-II). Fibromyalgia was defined as the sum of the z scores for muscle pain, muscle tension (FF), and somatic muscular (HAMA), whereas FibroFatigue is defined as the sum of all fatigue and fibromyalgia symptoms. Cognitive impairments include: concentration disorders, memory disturbances (both FF items), and intellectual impairments (HAMA). Insomnia was the sum of: the sleep disorder items of the FF, HAMA and BDI rating scales. The severity of autonomic symptoms was conceptualized as a z composite score of the autonomic disturbance items of the FF and HAMA scales.

Tobacco use disorder (TUD) was diagnosed according to DSM-IV-TR criteria. Body mass index (BMI) was computed using body weight in kilograms (kg) / length in meters (m)^2^.

### Measurements

Fasting venous blood samples were taken from the participants between 8.00 a.m. and 9.00 a.m. and collected into plain and EDTA tubes. Leukocyte and erythrocyte counts, packed cell volume (PCV), hemoglobin concentration, and hematimetric indices, namely MCV (mean corpuscular volume), MCH (mean corpuscular hemoglobin), MCHC (mean corpuscular hemoglobin concentration) and RDW (red blood cell distribution width) were measured by using the five-part differential Mindray^®^ BC-5000 hematology analyzer (Mindray Medical Electronics Co., Shenzhen, China). Samples were aliquoted and stored at − 80 C before assay. After separation, the sera were distributed into three new Eppendorf^®^ tubes for further analysis. Serum sodium, potassium, copper, and zinc were measured spectrophotometrically using kits supplied by Spectrum Diagnostics Co. (Cairo, Egypt). Glucose, albumin, total serum protein (TSP), urea, and creatinine were measured spectrophotometrically by kits supplied by Biolabo^®^ (Maizy, France). Calcium and magnesium levels in serum were measured spectrophotometrically using kits supplied by Spinreact^®^ (Barcelona, Spain). The C-Reactive Protein (CRP) latex slide test (Spinreact^®^, Barcelona, Spain) was used for CRP assays in human serum. The serum titer of CRP is the reciprocal of the maximum dilution exhibiting a positive response multiplied by a positive control concentration. The CRP concentration in the patient sample is calculated as follows: 6 x CRP Titer = mg/L. The total globulin fraction is calculated by subtracting albumin from total protein. The estimated GFR (eGFR) was calculated by using the Modification of Diet in Renal Disease Study equation (Levey et al., 2007) as: eGFR =175 x (S.Cr)^-1.154^ x (Age)^-0.203^ x 0.742 [if female] x 1.212 [if Black].

## Statistical Analysis

The Kolmogorov-Smirnov test was used to examine the normality of distributions. Using Pearson’s product moment correlation coefficients, correlations between variables were analyzed. Analysis of variance (ANOVA) was used to examine the between-group differences in scale variables. At a significance level of p < 0.05, pairwise comparisons of group means were conducted to identify differences among the three study groups. In addition, the false discovery rate (FDR) p-value was employed to correct for multiple comparisons (Benjamini and Hochberg, 1995). Statistical associations between categorical variables were evaluated by contingency table (χ^2^-test) analysis. We used multiple regression analysis (manual method) to assess the most significant predictors of the clinical rating scale scores using biomarkers as explanatory variables, while allowing for the effects of possible confounders (age, sex, BMI, TUD). In addition to the manual regression technique, we employed an automated technique with p-values of 0.05 for model entry and 0.10 for model removal. We calculated the model statistics (F, df, and p values) as well as the total variance explained (R^2^), standardised beta coefficients, t statistics, and exact p-values for each predictor. Additionally, the variance inflation factor and tolerance were evaluated in order to identify any collinearity or multicollinearity issues. Heteroskedasticity was determined with the help of the White and modified Breusch-Pagan homoscedasticity tests. As a method for feature reduction, principal component analysis (PCA) was used to create new PCs that reflect an underlying concept. To this end, the variance explained (VE) by the first principal component (PC) should be at least > 50%, and all variables should have high loadings on the first PC (i.e., > 0.66). In addition, the factoriability of the correlation matrix was evaluated using the Kaiser-Meyer-Olkin (KMO) Measure of Sampling Adequacy (values less than 0.5 indicate that corrective action is required, while values greater than 0.7 indicate a sampling that is more than adequate). Bartlett’s sphericity test is used to test the null hypothesis that the variables contained in the population correlation matrix are uncorrelated. In addition, we examined the anti-image correlation matrix as a measure of sampling adequacy. The visual binning method was employed to split the study group into subgroups based on continuous scores of kidney function. All preceding statistical analyses were conducted using IBM, SPSS for Windows version 28 (IBM-USA).

Partial least squares (PLS) analysis (Ringle, 2015) was used to determine the associations between biomarkers entered as input variables and symptom domain scores entered as output variables. Data were entered as either single indicators (all biomarkers) or a latent vector generated from the symptom dimensions. PLS path analysis was conducted using 5,000 bootstrap samples only when: a) the overall quality of the model as indicated by Standardized Root Mean Squared Error (SRMR) was adequate, namely SRMR < 0.08; b) the latent vector extracted from symptom dubdomain indicators has adequate congruence and reliability as indicated by average variance extracted (AVE) > 0.500, Cronbach’s alpha > 0.7, composite reliability > 0.7, and rho_A > 0.80; c) all indicators loaded highly (> 0.660) at p<0.001 on the latent vector; and d) construct cross-validated redundancies are adequate (Ringle, 2015). We employed complete bootstrapping (5,000 subsamples) and PLS path modeling to compute path coefficients with p-values and total and indirect effects (Luo et al., 2019). When using an effect size of 0.15 at p =0.05 (two-tailed) and power = 0.08, the estimated sample size should be 98, according to an a priori sample size calculation performed with G*Power 3.1.9.4 (multiple regression analysis with six covariates).

## Results

### Construction of renal functions PCs and ESRD subgroups

Using PCA we have constructed two PCs reflecting kidney functions and dialysis chracteristics. First, we were able to extract a validated PC from urea, creatinine, eGFR, TSP and albumin, labelled PC_Kidney (KMO=0.878, Bartlett’s test of sphericity χ2=368.872, df=10, p<0.001, VE=72.53%, all loadings > 0.783). Second, we extracted a PC from number dialyses/week, total number of dialyses and last day, labelled PC_dialysis (KMO=0.613, Bartlett’s test of sphericity χ2=113.336, df=3, p<0.001, VE=69.79%, all loadings > 0.774). Using a visual bining method, we divided the patients group into two subgroups using a cutoff score of −0.8 of the PC_kidney scores; the first subgroup (ESRD) comprised 43 paients and the second (extreme ESRD) 27 patients.

### Comparison of demographic and clinical data among the study groups

The sociodemographic and clinical data of the two ESRD patient groups and healthy controls are presented in **Table 1**. This table shows the differences in the PC_kidney scores among the three study groups. The data show results that are typical of the ESRD status, namely a significant increase in serum urea, creatinine, and lower eGFR, TSP and albumine in patients as compared with controls. Moreover, these changes were more pronounced in the extreme-ESRD subgroup. The number of dialysis sessions per week, total numberr, and the days after the last dialysis session were not significantly different between both ESRD groups. There were no significant differences in age, age at onset, sex ratio, BMI, TUD, residency, and employment between the three study groups. The family history of CKD was higher in the extreme-ESRD group. There were no signifcant differences in the drugs taken between the extreme-ESRD and ESRD subgroups (chlorpheniramine maleate, iron sucrose injections, sevelamer, and paracetamol). There were no differences in potassium, sodium, and Na/K ratio among both ESRD groups, but sodium was lower and potassium higher in patients than controls.

**Table 1.**
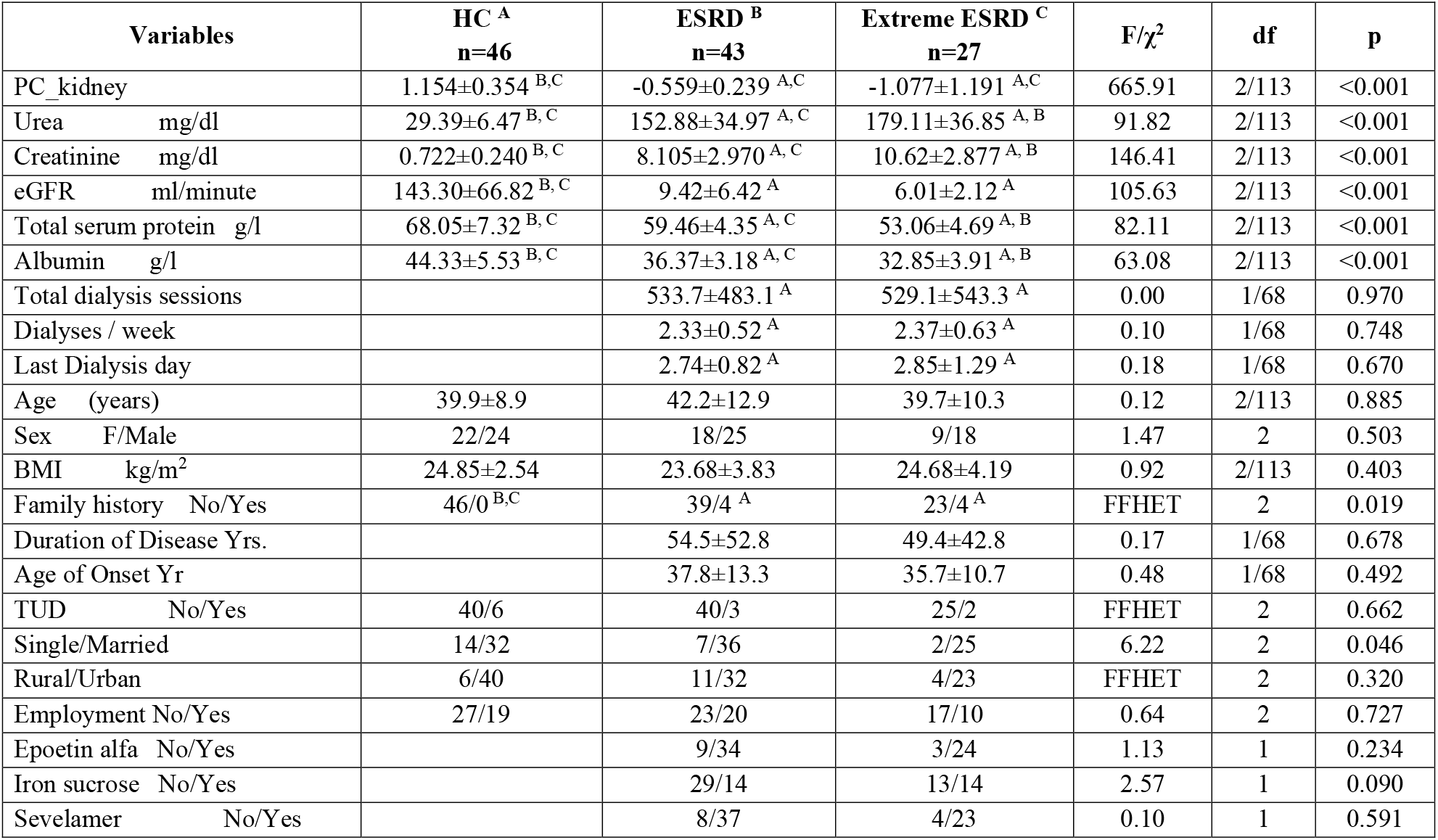
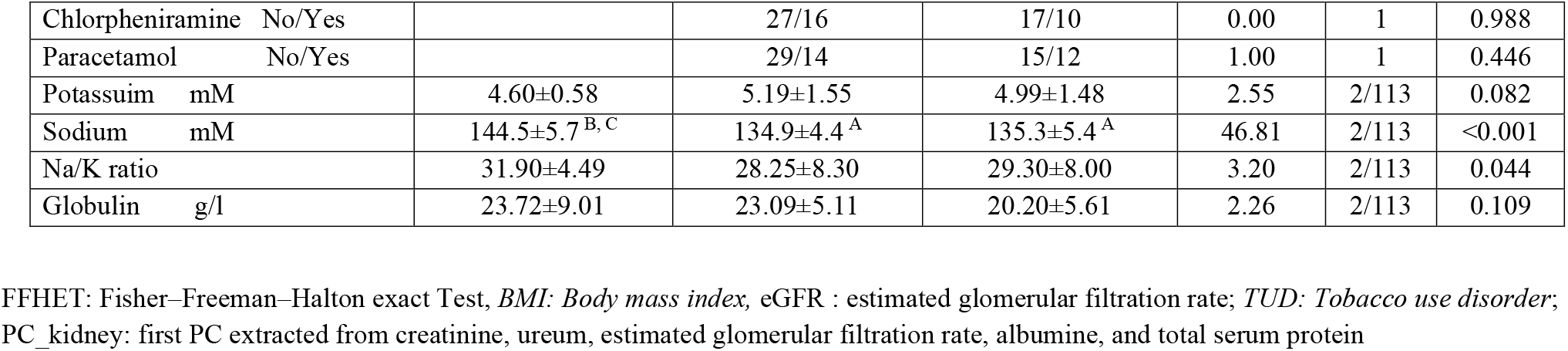
Demographic and biochemical data of healthy controls (HC) and end stage renal disease (ESRD) patients divided into those with and without extreme ESRD.

| Variables | HC <sup>A</sup><br>n=46 | ESRD <sup>B</sup><br>n=43 | Extreme ESRD <sup>C</sup><br>n=27 | F/$\chi^2$ | df | p |
| --- | --- | --- | --- | --- | --- | --- |
| PC_kidney | 1.154±0.354 <sup>B,C</sup> | -0.559±0.239 <sup>A,C</sup> | -1.077±1.191 <sup>A,C</sup> | 665.91 | 2/113 | <0.001 |
| Urea mg/dl | 29.39±6.47 <sup>B, C</sup> | 152.88±34.97 <sup>A, C</sup> | 179.11±36.85 <sup>A, B</sup> | 91.82 | 2/113 | <0.001 |
| Creatinine mg/dl | 0.722±0.240 <sup>B, C</sup> | 8.105±2.970 <sup>A, C</sup> | 10.62±2.877 <sup>A, B</sup> | 146.41 | 2/113 | <0.001 |
| eGFR ml/minute | 143.30±66.82 <sup>B, C</sup> | 9.42±6.42 <sup>A</sup> | 6.01±2.12 <sup>A</sup> | 105.63 | 2/113 | <0.001 |
| Total serum protein g/l | 68.05±7.32 <sup>B, C</sup> | 59.46±4.35 <sup>A, C</sup> | 53.06±4.69 <sup>A, B</sup> | 82.11 | 2/113 | <0.001 |
| Albumin g/l | 44.33±5.53 <sup>B, C</sup> | 36.37±3.18 <sup>A, C</sup> | 32.85±3.91 <sup>A, B</sup> | 63.08 | 2/113 | <0.001 |
| Total dialysis sessions |  | 533.7±483.1 <sup>A</sup> | 529.1±543.3 <sup>A</sup> | 0.00 | 1/68 | 0.970 |
| Dialyses / week |  | 2.33±0.52 <sup>A</sup> | 2.37±0.63 <sup>A</sup> | 0.10 | 1/68 | 0.748 |
| Last Dialysis day |  | 2.74±0.82 <sup>A</sup> | 2.85±1.29 <sup>A</sup> | 0.18 | 1/68 | 0.670 |
| Age (years) | 39.9±8.9 | 42.2±12.9 | 39.7±10.3 | 0.12 | 2/113 | 0.885 |
| Sex F/Male | 22/24 | 18/25 | 9/18 | 1.47 | 2 | 0.503 |
| BMI kg/m<sup>2</sup> | 24.85±2.54 | 23.68±3.83 | 24.68±4.19 | 0.92 | 2/113 | 0.403 |
| Family history No/Yes | 46/0 <sup>B,C</sup> | 39/4 <sup>A</sup> | 23/4 <sup>A</sup> | FFHET | 2 | 0.019 |
| Duration of Disease Yrs. |  | 54.5±52.8 | 49.4±42.8 | 0.17 | 1/68 | 0.678 |
| Age of Onset Yr |  | 37.8±13.3 | 35.7±10.7 | 0.48 | 1/68 | 0.492 |
| TUD No/Yes | 40/6 | 40/3 | 25/2 | FFHET | 2 | 0.662 |
| Single/Married | 14/32 | 7/36 | 2/25 | 6.22 | 2 | 0.046 |
| Rural/Urban | 6/40 | 11/32 | 4/23 | FFHET | 2 | 0.320 |
| Employment No/Yes | 27/19 | 23/20 | 17/10 | 0.64 | 2 | 0.727 |
| Epoetin alfa No/Yes |  | 9/34 | 3/24 | 1.13 | 1 | 0.234 |
| Iron sucrose No/Yes |  | 29/14 | 13/14 | 2.57 | 1 | 0.090 |
| Sevelamer No/Yes |  | 8/37 | 4/23 | 0.10 | 1 | 0.591 |
| Chlorpheniramine No/Yes |  | 27/16 | 17/10 | 0.00 | 1 | 0.988 |
| Paracetamol No/Yes |  | 29/14 | 15/12 | 1.00 | 1 | 0.446 |
| Potassium mM | 4.60±0.58 | 5.19±1.55 | 4.99±1.48 | 2.55 | 2/113 | 0.082 |
| Sodium mM | 144.5±5.7<sup>B, C</sup> | 134.9±4.4<sup>A</sup> | 135.3±5.4<sup>A</sup> | 46.81 | 2/113 | <0.001 |
| Na/K ratio | 31.90±4.49 | 28.25±8.30 | 29.30±8.00 | 3.20 | 2/113 | 0.044 |
| Globulin g/l | 23.72±9.01 | 23.09±5.11 | 20.20±5.61 | 2.26 | 2/113 | 0.109 |

FFHET: Fisher–Freeman–Halton exact Test, *BMI*: *Body mass index*, eGFR : estimated glomerular filtration rate; *TUD*: *Tobacco use disorder*; PC\_kidney: first PC extracted from creatinine, ureum, estimated glomerular filtration rate, albumine, and total serum protein

### Differences in hematological biomarkers and electrolytes in the study groups

The biomarker levels in both patient groups and the healthy controls are presented in **Table 2**. The number of WBCs was lower in patients than controls and CRP was increased in the patients. Number RBCs PCV, hemoglobin and MCHC were significantly lower in patients than controls. We were able to extract one validated PC from these 4 RBC measuremets, labeled PC_RBC (KMO=0.537, Bartlett’s test of sphericity χ2=928.937, df=6, p<0.001, VE=80.83%, all loadings > 0.723). PC_RBC was significantly lower in patients than controls. We were also able to extract one validated PC from MCV, MCH and RDW-CV, labeled PC_RBCindices (KMO=0.574, Bartlett’s test of sphericity χ2=174.739, df=3, p<0.001, VE=70.99%, all loadings > 0.705). PC_RBCindices and MCH were lower in the extreme-ESRD group than in controls. MCV was higher in ESRD patients than controls, and RDW-CV higher in patients than controls. No significant differences were established in serum magnesium among the study groups. Calcium and zinc (both lowered) and copper (increased) were significantly different between patients and controls.

**Table 2.** Measurements of biomarkers in the healthy controls (HC) and end stage renal disease (ESRD) patients divided into those with and without extreme ESRD.

| Variables | Control <sup>A</sup><br>n=46 | ESRD <sup>B</sup><br>n=43 | Extreme ESRD <sup>C</sup><br>n=27 | F | df | p |
| --- | --- | --- | --- | --- | --- | --- |
| WBC 10<sup>9</sup>/L | 6.88±2.18 <sup>B,C</sup> | 5.96±2.11 <sup>A</sup> | 5.57±1.86 <sup>A</sup> | 3.95 | 2/113 | 0.022 |
| CRP mg/l | 0.84±0.53 <sup>B,C</sup> | 9.31±17.70 <sup>A</sup> | 4.64±6.34 <sup>A</sup> | 23.21 | 2/113 | <0.001 |
| RBCs million/μL | 5.11±0.65 <sup>B,C</sup> | 3.61±0.66 <sup>A</sup> | 3.73±0.65 <sup>A</sup> | 69.54 | 2/113 | <0.001 |
| PCV % | 40.81±4.64 <sup>B,C</sup> | 30.61±5.05 <sup>A</sup> | 30.44±5.15 <sup>A</sup> | 61.34 | 2/113 | <0.001 |
| Hemoglobin g/dl | 13.52±1.31 <sup>B,C</sup> | 9.13±1.53 <sup>A</sup> | 9.02±1.48 <sup>A</sup> | 132.68 | 2/113 | <0.001 |
| MCHC g/dl | 33.22±1.53 <sup>B,C</sup> | 29.91±0.98 <sup>A</sup> | 29.68±1.05 <sup>A</sup> | 104.46 | 2/113 | <0.001 |
| PC_RBC | 1.053±0.472 <sup>B,C</sup> | -0.675±0.536 <sup>A</sup> | -0.693±0.500 <sup>A</sup> | 165.04 | 2/113 | <0.001 |
| MCV fL | 80.40±7.28 <sup>B</sup> | 85.26±9.25 <sup>A</sup> | 82.45±11.09 | 3.26 | 2/113 | 0.042 |
| MCH pg | 26.41±3.19 <sup>C</sup> | 25.48±2.67 | 24.48±3.41 <sup>A</sup> | 3.44 | 2/113 | 0.035 |
| RDW-CV % | 13.51±2.07 <sup>B,C</sup> | 16.99±1.76 <sup>A</sup> | 17.69±2.42 <sup>A</sup> | 47.26 | 2/113 | <0.001 |
| PC_RBCindices | 0.287±0.835 <sup>C</sup> | -0.054±0.949 | -0.402±1.201 <sup>A</sup> | 4.38 | 2/113 | 0.015 |
| Magnesium mM | 0.777±0.169 | 0.838±0.165 | 0.791±0.176 | 1.55 | 2/113 | 0.217 |
| Calcium mM | 2.270±0.162 <sup>B,C</sup> | 2.033±0.316 <sup>A</sup> | 1.920±0.260 <sup>A</sup> | 18.99 | 2/113 | <0.001 |
| Copper mg/l | 0.762±0.278 <sup>B,C</sup> | 1.157±0.284 <sup>A</sup> | 1.166±0.302 <sup>A</sup> | 26.95 | 2/113 | <0.001 |
| Zinc mg/l | 0.814±0.146 <sup>B,C</sup> | 0.636±0.197 <sup>A</sup> | 0.722±0.211 <sup>A</sup> | 10.53 | 2/113 | <0.001 |

CRP: C-reactive protein; MCH: mean corpuscular hemoglobin; MCHC: mean corpuscular hemoglobin concentration; MCV: mean corpuscular volume; RBCs: red blood cells count; PC\_RBCs: principal component of number RBCs, PCV, hemoglobin and MCHC; PC\_RBCindices: principal component of MCV, MCH and RDW-CV; PCV: packed cell volume; RDW-CV: coefficient of variance of RBCs distribution width; WBCs: white blood cells count

### Differences in neuropsychiatric scores among the study groups

The scores of psychiatric subdomains in the healthy controls (HC) and both ESRD subgroups are presented in **Table 3**. All subdomain scores were sigificantly higher in patients than controls. There were significant differences between the three groups in pure physiosomatic and fibromyalgia and fibro-fatigue symptoms with increasing scores from controls to ESRD to extreme-ESRD.

**Table 3.**
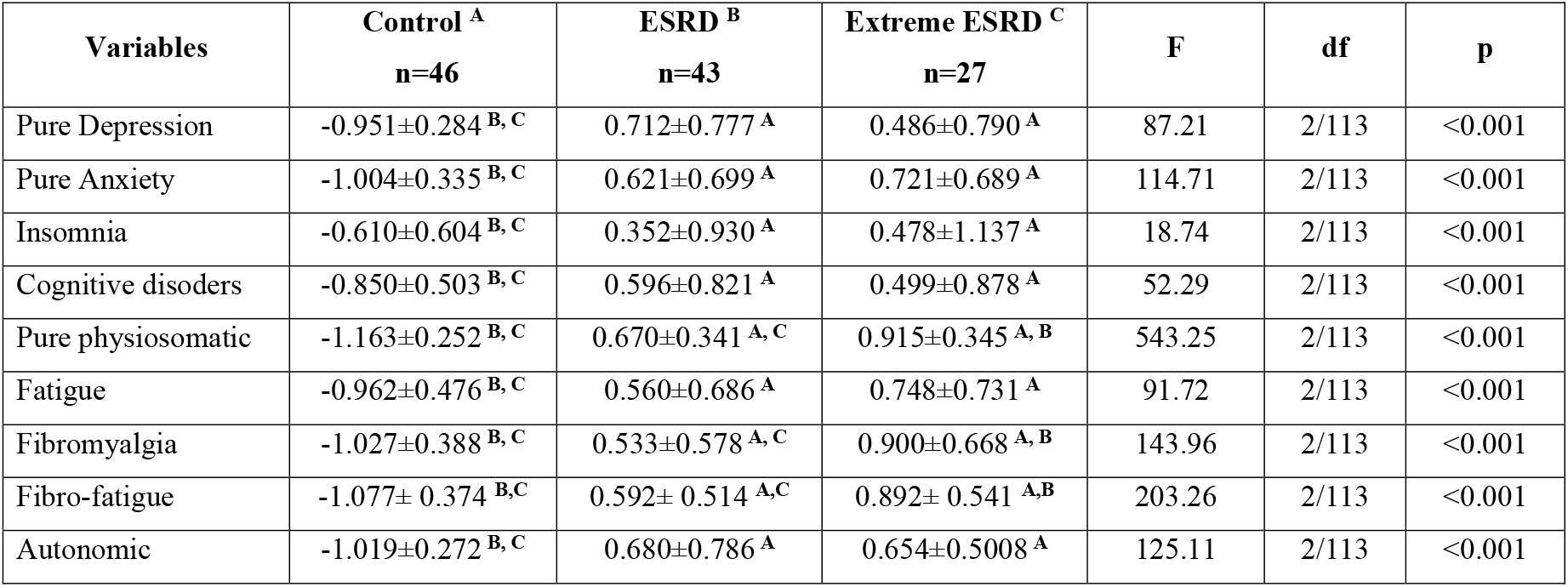
Measurements of neuropsychiatric subdomain scores in healthy controls (HC) and end-stage renal disease (ESRD) patients divided into those with and without extreme ESRD.

### Prediction of RBCs parameters

**Table 4** shows the results of multiple regression analyses with PC_RBC and PC_RBCindices as dependent variables and PC_kidney, PC_dialysis, and biomarkers as explanatory variables. The first regression shows that 61.7% of the variance in PC_RBC was explained by PC_kidney and calcium (both positively), and PC_dialysis and CRP (both inversely associated). **Figure 1** displays the partial regression of PC_RBCs on PC_kidney, while **Figure 2** presents the partial regression of PC_RBC on the PC_dialysis. Regression #2 reveals that 41.1% of the variance in the PCV could be explained by calcium and PC_Kidney (both positively) associated, and PC_dialysis (inversely associated). In Regression #3, a large part of the variance (59.7%) in hemoglobin was explained by PC_kidney, calcium, and BMI (positively associated) along with PC_dialysis and CRP (inversely associated). Regression #4 shows that 10.4% of the variance in PC_RBCindices could be explained by PC_kidney and BMI.

**Table 4.** Results of multiple regression analyses with red blood cell (RBC) features as dependent variables and renal kidney and biomarkers as explanatory variables

| Dependent Variables | Explanatory Variables | $\beta$ | t | p | F model | df | p | R<sup>2</sup> |
| --- | --- | --- | --- | --- | --- | --- | --- | --- |
| **#1. PC_RBCs** | **Model #1** |  |  |  | 44.27 | 4/110 | <0.001 | 0.617 |
|  | PC_kidney | 0.323 | 3.89 | <0.001 |  |  |  |  |
|  | PC_dialysis | -0.289 | -3.34 | 0.001 |  |  |  |  |
|  | Calcium | 0.223 | 3.39 | <0.001 |  |  |  |  |
|  | CRP | -0.180 | -2.57 | 0.012 |  |  |  |  |
| **#2. PCV** | **Model #2** |  |  |  | 25.79 | 3/111 | <0.001 | 0.411 |
|  | PC_dialysis | -0.314 | -3.18 | 0.002 |  |  |  |  |
|  | Calcium | 0.264 | 3.26 | 0.001 |  |  |  |  |
|  | PC_kidney | 0.214 | 2.10 | 0.038 |  |  |  |  |
| **#3. Hemoglobin** | **Model #3** |  |  |  | 32.33 | 5/109 | <0.001 | 0.597 |
|  | PC_kidney | 0.301 | 3.52 | <0.001 |  |  |  |  |
|  | PC_dialysis | -0.276 | -3.09 | 0.003 |  |  |  |  |
|  | Calcium | 0.224 | 3.29 | 0.001 |  |  |  |  |
|  | CRP | -0.173 | -2.38 | 0.019 |  |  |  |  |
|  | BMI | 0.146 | 2.37 | 0.020 |  |  |  |  |
| **#4. PC_RBCindices** | **Model #5** |  |  |  | 6.47 | 2/112 | 0.002 | 0.104 |
|  | PC_kidney | 0.239 | 2.66 | 0.009 |  |  |  |  |
|  | BMI | 0.204 | 2.28 | 0.024 |  |  |  |  |

BMI: Body mass index, PCV: packed cell volume; PC\_dialyses: principal component extracted of dialysis parameters; PC\_kidney: principal component extracted from kidney parameters; PC\_RBCs: principal component extracted from number of RBC, packed cell volume, hemoglobin and mean corpuscular hemoglobin concentration; RBCs: red blood cells count; PC\_RBCindices: principal component extracted from mean corpuscular hemoglobin, mean corpuscular volume and red cell distribution width; CRP: C-reactive protein.

**Figure 1.**
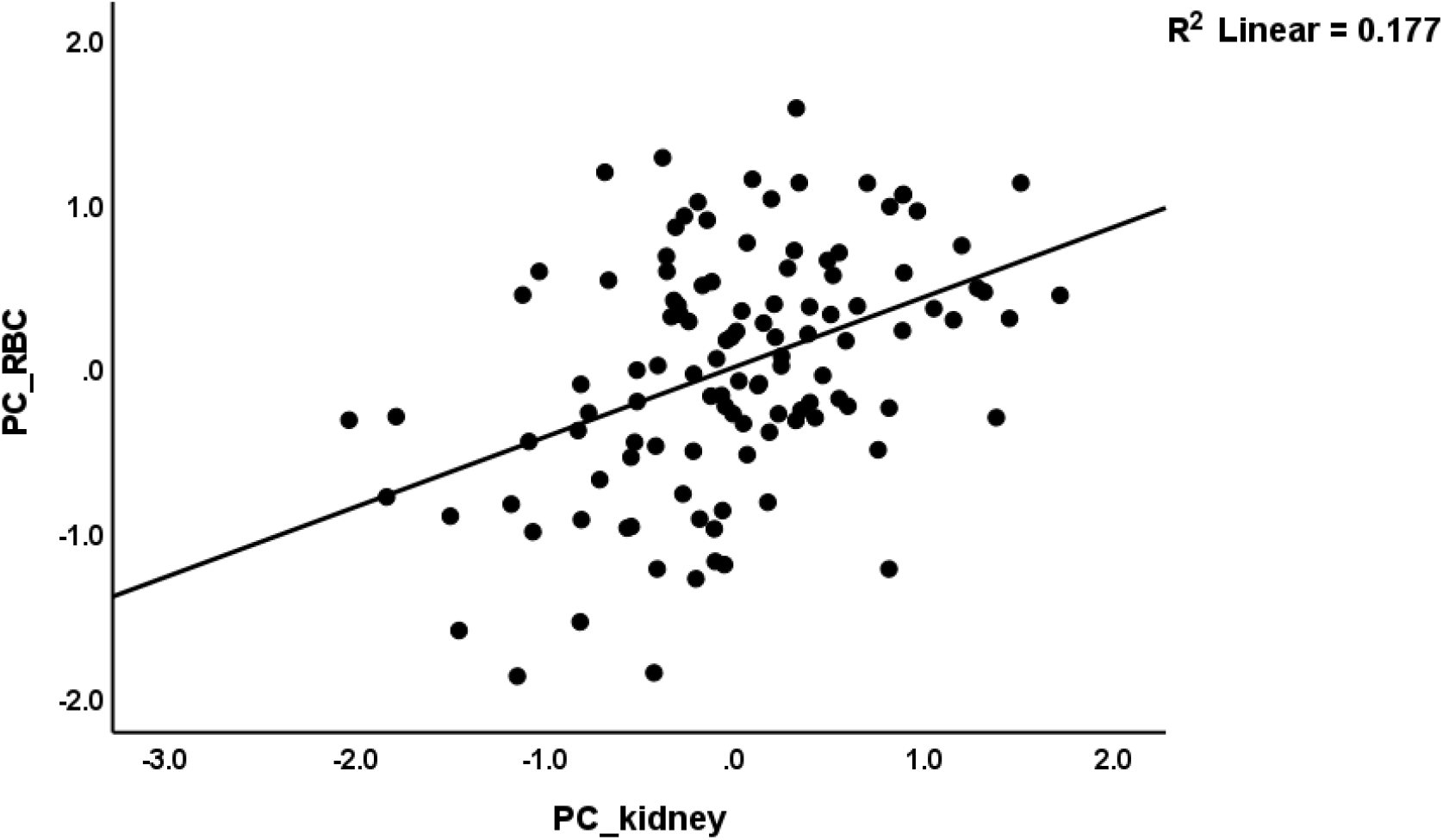
Partial regression of the first principal component (PC) extracted from number of red blood cells, hematocrit, hemoglobin, and mean corpuscular hemoglobin concentration) on the first PC extracted from creatinine, ureum, estimated glomerular filtration rate, albumine, and total serum protein (PC_kidney).

**Figure 2.**
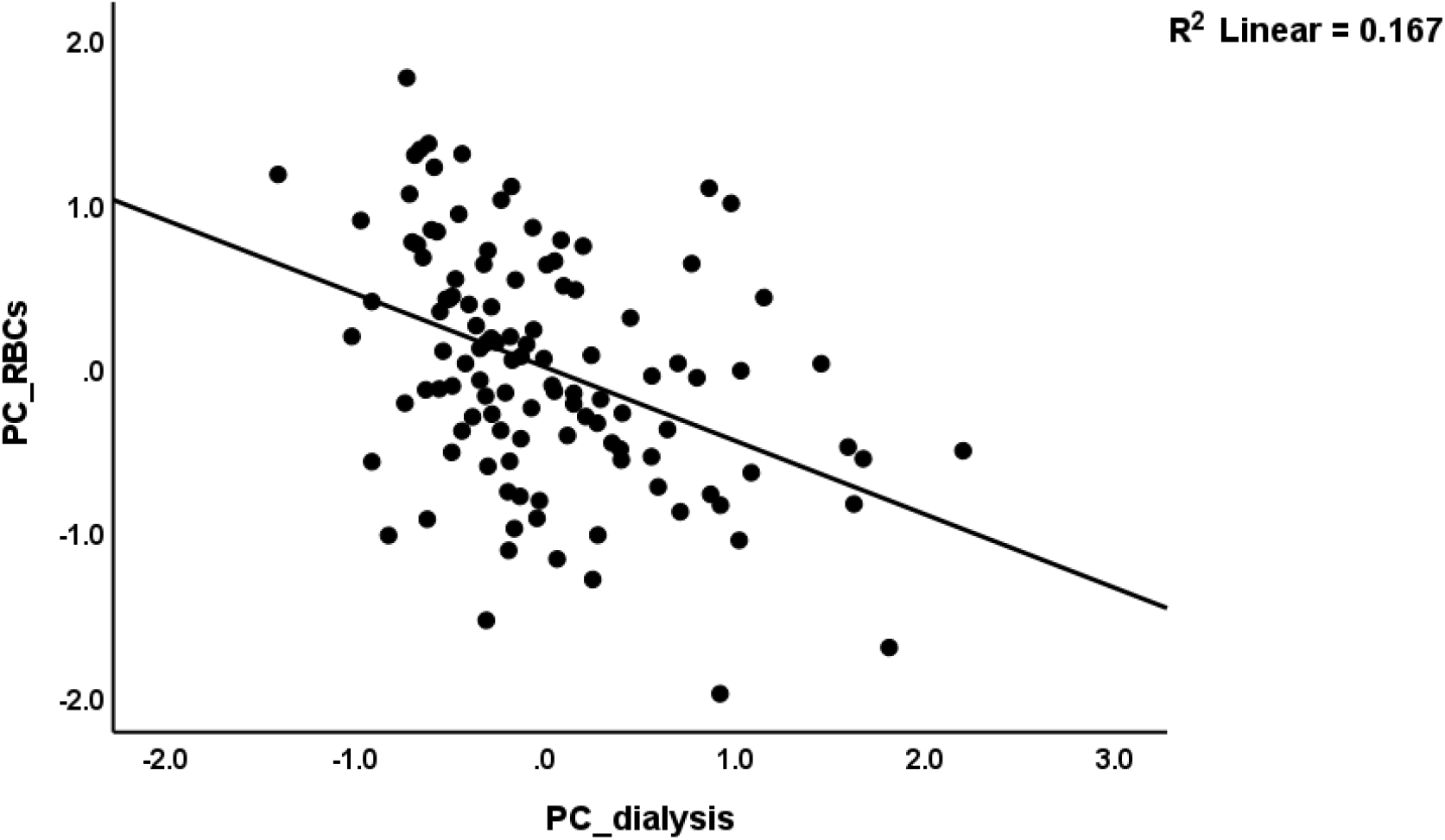
Partial regression of the first principal component (PC) extracted from number of red blood cells, hematocrit, hemoglobin, and mean corpuscular hemoglobin concentration, on the first PC extracted from dialysis indicators (e.g. total number, weekly number)

### Prediction of neuropsychiatric symptom domains

**Table 5** shows the results of different stepwise multiple regression analyses with neuropsychiatric symptom scores as dependent variables and biomarkers and clinical characteristics as independent variables. Regression #1 shows that 61.0% of the variance in the pure depression score was explained by PC_RBC, zinc, and PC_kidney (inversely associated), CRP (positively associated) and male sex. **Figure 3** shows the partial regression of pure depressive symptoms on PC_RBC. Regression #2 shows that the model explained 67.9% of the variance in the pure anxiety score using CRP and copper (positively associated), age, PC_kidney, PC_RBCs, and zinc (negatively associated) as explanatory variables. Regression #3 shows that 82.1% of the variance in the pure physiosomatic symptoms score was explained by the copper and CRP (positively) and PC_RBC, PC-kidney, and calcium (inversely associated). **Figures 4** and **5** show the partial regressions of the pure physiosomatic domain scores on PC_RBCs and PC_kidney, respectively. Regression #4 shows that 24.4% of the variance in insomnia was explained by copper (positively associated) and PC_RBC (inversely associated). Regression #5 shows that a considerable part of the variance in cognition score (44.8%) was explained by the regression on copper and CRP (positively associated) and PC_RBC (inversely associated). Regression #6 shows that 58.6% of the variance in fatigue was explained by PC_RBCs (inversely associated). Regression #7 shows that a significant part of the fibromyalgia variance (62.7%) was explained by PC_kidney and PC_RBCs (inversely associated) and copper (positively associated). Regression #8 shows that 67.3% of the variance in the autonomic symptom scores was explained by copper, CRP, and sex (positively associated), and PC_kidney and PC_RBCs (inversely associated).

**Table 5.** Results of multiple regression analyses with neuropsychiatric subdomain scores as dependent variables

| Dependent Variables | Explanatory Variables | $\beta$ | t | p | F model | df | p | R<sup>2</sup> |
| --- | --- | --- | --- | --- | --- | --- | --- | --- |
| **#1. Pure depression** | **Model #1** |  |  |  | 34.09 | 5/109 | <0.001 | 0.610 |
|  | PC_RBC | -0.294 | -3.32 | 0.001 |  |  |  |  |
|  | CRP | 0.340 | 4.84 | <0.001 |  |  |  |  |
|  | Zinc | -0.203 | -3.26 | 0.001 |  |  |  |  |
|  | PC_kidney | -0.205 | -2.47 | 0.015 |  |  |  |  |
|  | Sex | 0.132 | 2.17 | 0.032 |  |  |  |  |
| **#2. Pure Anxiety** | **Model #2** |  |  |  | 38.06 | 6/108 | <0.001 | 0.679 |
|  | PC_RBCs | -0.237 | -2.90 | 0.005 |  |  |  |  |
|  | CRP | 0.338 | 5.29 | <0.001 |  |  |  |  |
|  | PC_kidney | -0.268 | -3.45 | <0.001 |  |  |  |  |
|  | Zinc | -0.153 | -2.65 | 0.009 |  |  |  |  |
|  | Copper | 0.166 | 2.67 | 0.009 |  |  |  |  |
|  | Age | -0.115 | -2.09 | 0.039 |  |  |  |  |
| **#3. Pure**<br>**physiosomatic** | **Model #3** |  |  |  | 99.87 | 5/109 | <0.001 | 0.821 |
|  | PC_RBCs | -0.430 | -6.85 | <0.001 |  |  |  |  |
|  | PC_kidney | -0.287 | -4.91 | <0.001 |  |  |  |  |
|  | Copper | 0.202 | 4.45 | <0.001 |  |  |  |  |
|  | CRP | 0.145 | 3.06 | 0.003 |  |  |  |  |
|  | Calcium | -0.109 | -2.308 | 0.023 |  |  |  |  |
| **#4. Insomnia** | **Model #4** |  |  |  | 18.05 | 2/112 | <0.001 | 0.244 |
|  | PC_RBC | -0.299 | -3.37 | 0.001 |  |  |  |  |
|  | Copper | 0.296 | 3.33 | 0.001 |  |  |  |  |
| **#5. Cognition** | Model #5 |  |  |  | 30.00 | 3/111 | <0.001 | 0.448 |
|  | PC_RBC | -0.423 | -4.86 | <0.001 |  |  |  |  |
|  | Copper | 0.221 | 2.89 | 0.005 |  |  |  |  |
|  | CRP | 0.213 | 2.61 | 0.010 |  |  |  |  |
| **#6. Fatigue** | **Model #6** |  |  |  | 159.82 | 1/113 | <0.001 | 0.586 |
|  | PC_RBCs | -0.765 | -12.642 | <0.001 |  |  |  |  |
| **#7. Fibromyalgia** | **Model #7** |  |  |  | 62.21 | 3/111 | <0.001 | 0.627 |
|  | PC_RBCs | -0.417 | -5.195 | <0.001 |  |  |  |  |
|  | Copper | 0.247 | 3.818 | <0.001 |  |  |  |  |
|  | PC_kidney | -0.293 | -3.562 | <0.001 |  |  |  |  |
| **#8. Autonomic** | **Model #8** |  |  |  | 44.88 | 5/109 | <0.001 | 0.673 |
|  | PC_RBCs | -0.416 | -5.167 | <0.001 |  |  |  |  |
|  | Copper | 0.245 | 4.000 | <0.001 |  |  |  |  |
|  | CRP | 0.216 | 3.364 | 0.001 |  |  |  |  |
|  | PC_kidney | -0.163 | -2.073 | 0.041 |  |  |  |  |
|  | Sex | 0.115 | 2.069 | 0.041 |  |  |  |  |

BMI: Body mass index, PCV: packed cell volume; PC\_dialyses: principal components of dialysis parameters; PC\_kidney: principal component extracted from kidney parameters; PC\_RBCs: principal component extracted from number of RBC, packed cell volume, hemoglobin and mean corpuscular hemoglobin concentration; RBCs: red blood cells count; CRP: C-reactive protein.

**Figure 3.**
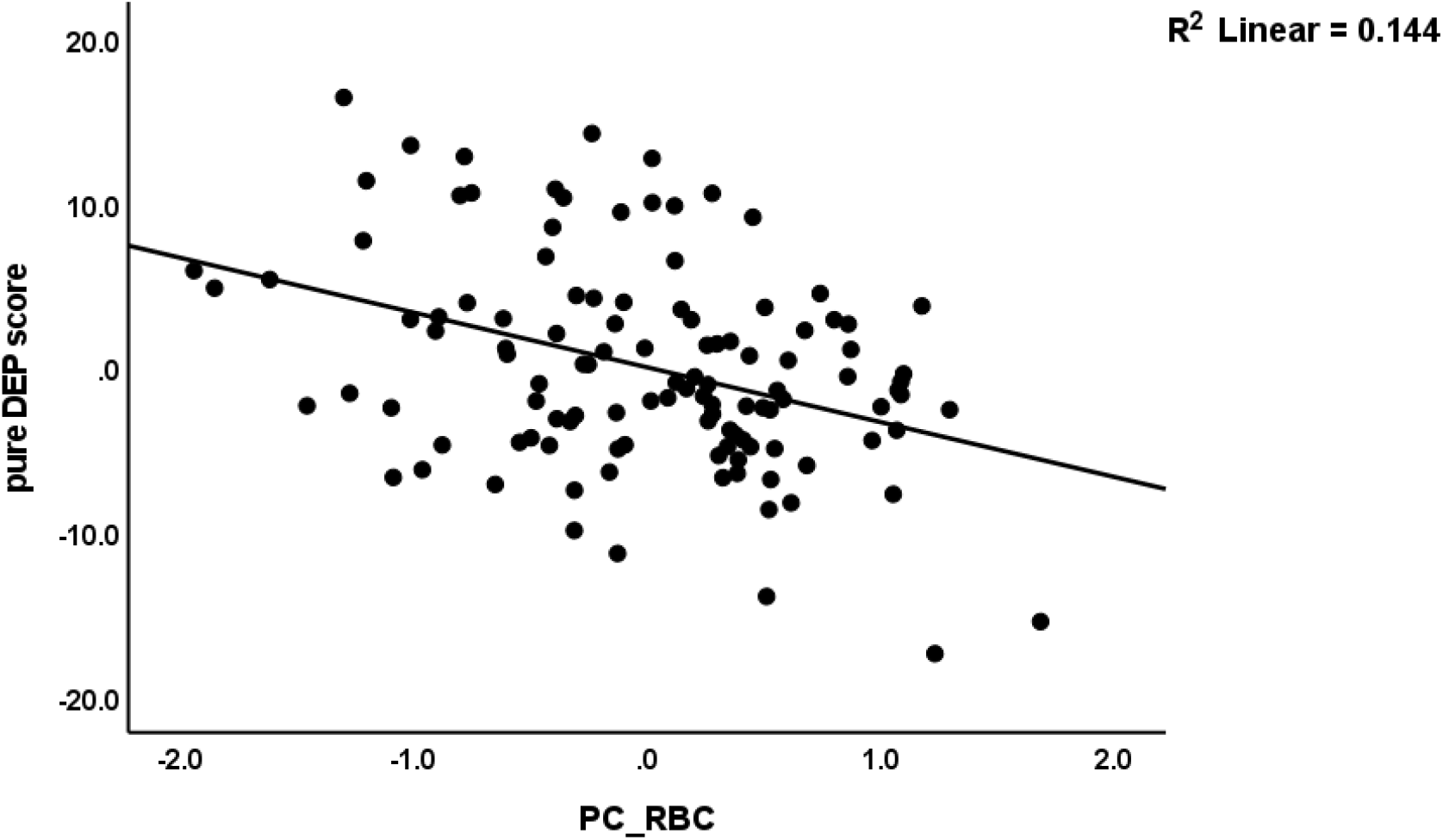
Partial regression of the pure depressive symptom (DEP) score on the first principal component extracted from red blood cell (RBC) features (number of RBC, hematocrit, hemoglobin, mean corpuscular hemoglobin concentration).

**Figure 4.**
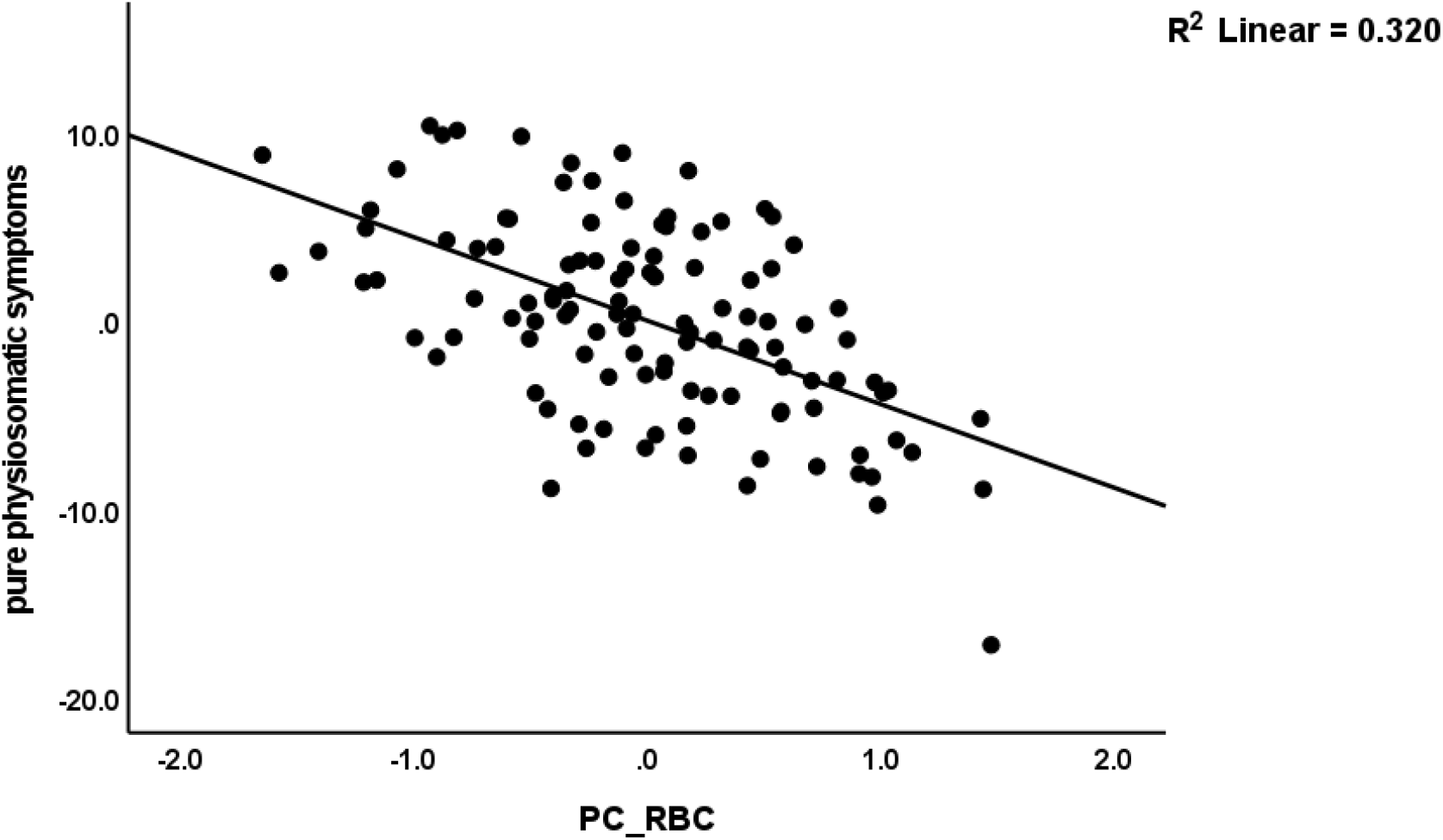
Partial regression of the pure physiosomatic symptoms on the first principal component extracted from number of red blood cells (RBC), hematocrit, hemoglobin, and mean corpuscular hemoglobin concentration)

**Figure 5.**
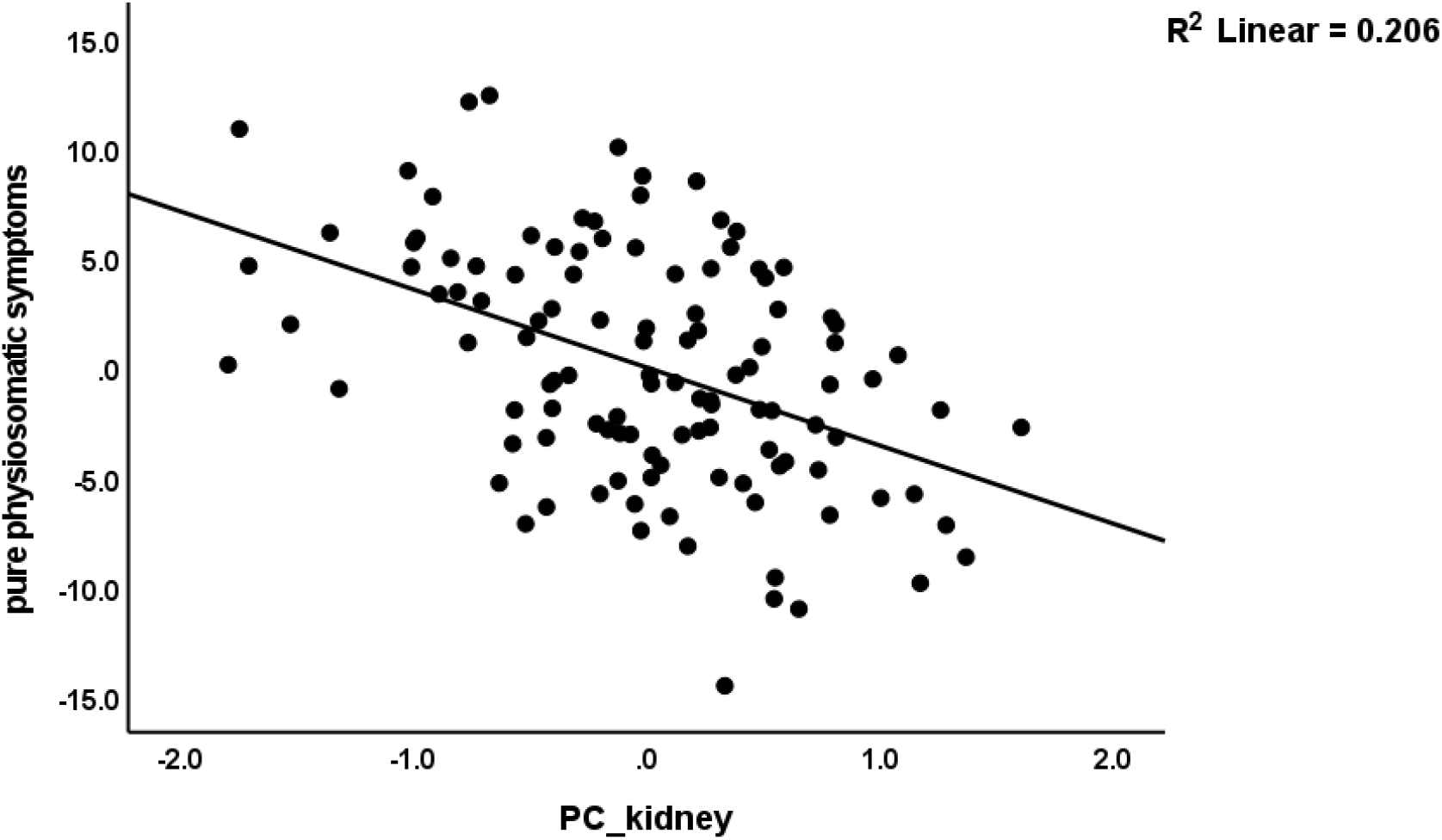
Partial regression of the pure physiosomatic symptoms on the first principal component extracted from creatinine, ureum, estimated glomerular filtration rate, albumine, and total serum protein (PC_kidney).

### Results of PLS path and PLS predict analysis

Figure 6. shows the final PLS model obtained after feature selection, prediction-oriented segmentation with multi-group analysis. The neuropsychiatric symptom domains were entered as a latent vector (LV) extracted from pure anxiety, pure depression, pure physiosomatic, autonomic, cognition, fatigue and fibromyalgia symptoms (the loading of insomnia was 0.603, and thus this doman was not included), LV_RBC was extracted from RBC, hemoglobin, PCV and MCHC, LV_kidney was extracted from albumin, TSP, eGFR, creatinine and ureum, and LV_dialysis was extracted from the three dialysis indicators. Copper, zinc an CRP were entered as single indicators. The model fit quality data are adequate with SRMR = 0.043, and the extracted LVs showed AVE values > 0.698, Cronbach’s alpha > 0.781, composite reliability > 0.873, and rho A > 0.809. PLS Blinfolding demonstrated that the construct crossvalidated redundancies of LVs were all > 0.360. PLS Predict showed that all Q2 values were positive incdicating sifficient replicability of the model. Complete PLS analysis, performed using 5,000 bootstraps, showed that 85.0% of the variance in the phenome was explained by LV_RBC, LV_kidney, copper, zinc and CRP. LV_kidney explained 64.6% and 27.3% in LV_RBC and copper, respectively. LV_dialysis explained 11.7% and 29.9% of the variance in zinc and CRP, respectively. There were significant specific indirect effects of LV_kidney on the phenenome that were mediated by copper (t=2.75, p=0.003) and RBC (t=3.84, p<0.001), and from LV_dialysis that were mediated by CRP (t=2.71, p=0.003).

**Figure 6.**
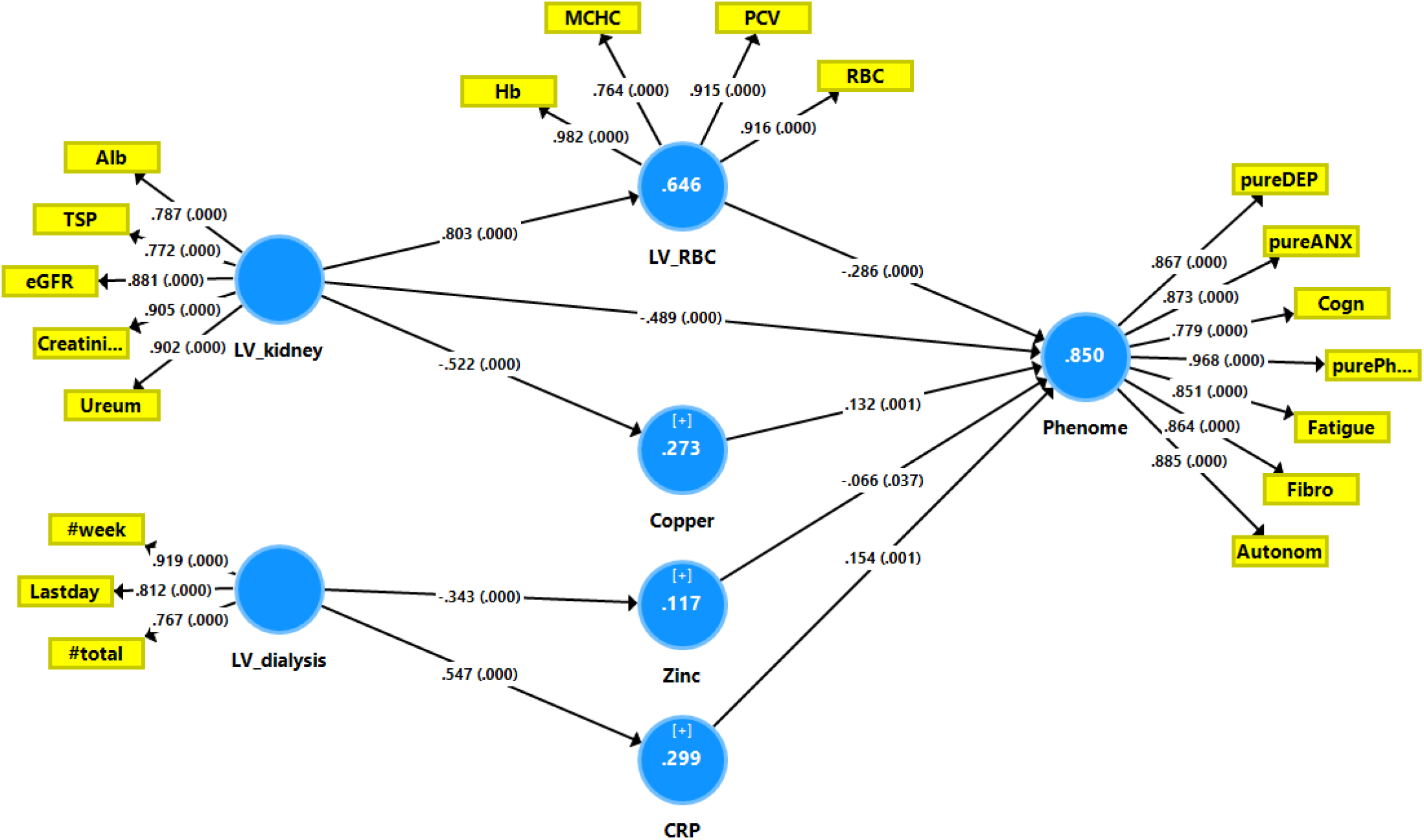
Results of Partial Least Squares analysis. The neuropsychiatric symptom domains were entered as a latent vector (LV) extracted from pure anxiety (pureANX), pure depression (pureDEP), pure physiosomatic (purePh), autonomic (autonom), cognition (cogn), fatigue and fibromyalgia (Fibro) symptoms. LV_RBC was extracted from red blood cell number (RBC), hemoglobin (Hb), packed cell volume (PCV) and mean corpuscular hemoglobin concentrations (MCHC). LV_kidney was extracted from albumin, total serum protein (TSP), estimated glomerular filtration rate (eGFR), creatinine (creatin) and ureum. LV_dialysis was extracted from the three dialysis indicators, namely total number of dialyses (#total), weekly number of dialyses (#week) and time lag to last day of dialysis (Lastday). CRP: C-reactive protein. Shown are path coefficients (with p value) of the inner model, and loadings (with p values) of the outer model. White figures in blue circles denote explained variance.

## Discussion

### Erythron biomarkers of ESRD

The first important finding of the current study is that ESRD is accompanied by significant changes in the erythron, including the number of red blood cells, PCV, haemoglobin, MCV, and RDW. The overall results indicate chronic illness-related anaemia in ESRD. In addition, these changes and the decreased PCV and haemoglobin levels are largely attributable to the combined effects of declining kidney function tests and an increasing number of dialysis sessions. Previous research demonstrated that, in the dialysis group, haemoglobin levels, RBC count, MCV, and MCHC values were significantly lower than in the control group (Belo et al., 2019). Increased baseline RDW was associated with a decline in residual renal functions in continuous ambulatory peritoneal dialysis (CAPD) patients, indicating that RDW is a useful indicator for the risk of residual renal function decline (Li et al., 2022).

There are two primary reasons for these correlations. the accumulation of uremic toxins in the blood leads to low erythropoietin production by the kidney interstitial fibroblasts (peritubular cells) resulting in defective erythropoiesis due to a decrease in erythrocyte production (Macdougall, 2001; Ratcliffe, 1993). Approximately fifty percent of ESRD patients in the United States have anaemia due to erythropoietin deficiency (McClellan et al., 2004). The second theory posits that the changes in RBC fragility and properties are caused by an increase in oxidative stress resulting from renal failure and renal damage (Rysz et al., 2020; Sangeetha Lakshmi et al., 2018; Song et al., 2020). OS, particularly H_2_O_2_, modifies the characteristics of RBC membranes and accelerates RBC clearance in the spleen (Nuhu and Bhandari, 2018; Srour et al., 2000). Increased RBC oxidative damage and reactive oxygen species (ROS) production as a result of iron deficiency anaemia in CKD result in a vicious cycle of RBC death, anaemia, and OS (Corrons et al., 1995; Nuhu and Bhandari, 2018; Srour et al., 2000). In addition, iron deficiency-related anaemia is accompanied by a decrease in oxygen partial pressure, which exacerbates OS by auto-oxidizing haemoglobin to met-hemoglobin (metHb) and producing O_2_ (Hébert et al., 2004; Nagababu et al., 2008). In addition, iron deficiency may influence the production of iron-containing endogenous antioxidant proteins, such as peroxidase and catalase, as well as selenium levels, thereby reducing glutathione peroxidase activity (Lazarte et al., 2015; Rifkind and Nagababu, 2013). Intravenous iron replenishment and antioxidants may improve the quality of life of CKD patients and reduce mortality (Steinmetz, 2012).

### Zinc, copper, calcium and CRP in ESRD

Compared to controls, ESRD patients exhibited altered copper, zinc, and calcium levels, as well as changes in electrolytes. In numerous studies, changes in trace elements in CKD/ESRD have been described (Ari et al., 2011; Hasanato, 2014; Navarro-Alarcon et al., 2006; Tonelli et al., 2009). Zinc plasma levels were significantly lower in CKD patients compared to controls (Tavares et al., 2021), and hemodialysis patients are at an increased risk of zinc deficiency (Ahmadipour et al., 2017; Gómez de Oña et al., 2016; Takahashi, 2022). Inflammation can also impact zinc levels (Maes, M. et al., 1997). Zinc supplementation may raise haemoglobin levels in dialysis-dependent ESRD patients (Fukushima et al., 2009; Kobayashi et al., 2015). The mechanism underlying the association between zinc and hemoglobin production involves a) the incorporation of zinc into the zinc-finger protein transcription factor (GATA-1) that regulates various genes involved in RBC synthesis (Katsumura et al., 2013), and 2) signaling cascades such as the growth hormone and insulin-like growth factor-1 pathway which influence RBC synthesis (Lytras and Tolis, 2007). Due to the fact that zinc is required for the activation of superoxide dismutase, a potent antioxidant enzyme (Prasad and Bao, 2019), a decrease in zinc levels may be accompanied by a decrease in radical scavenging defences. Zinc deficiency is associated with increased production of oxygen-free radicals, decreased immune functions, increased inflammation, delayed wound healing, and increased susceptibility to infection (Hirano et al., 2008; Prasad, 1985).

Zinc concentrations in hemodialysis patients may be negatively correlated with serum copper levels, which are frequently elevated in CKD (Almeida et al., 2020; Nishime et al., 2020). In addition, hemodialysis patients with anaemia have higher serum copper levels than those without anaemia, which may be due to copper’s detrimental effect on iron absorption and haemoglobin production (Zeraati et al., 2014). In patients undergoing peritoneal dialysis, elevated Cu/Zn ratios are associated with malnutrition, increased oxidative stress, inflammation, and a compromised immune status (Atakul et al., 2020). In addition, elevated copper levels (whether or not coupled with diminished zinc availability) may result in nephrotoxicity and tube necrosis (Niu et al., 2020).

Calcium-phosphate homeostasis disturbances are frequently observed in CKD and ESRD (Block et al., 2013), and in stages 3-4, reduced calcium levels are associated with worse outcomes (Lim et al., 2014). Hypocalcemia in chronic renal failure is caused by elevated serum phosphorus and diminished renal vitamin D production (Pazianas and Miller, 2021). The former induces hypocalcemia through complexation with serum calcium and deposition in bone and other tissues (Lim and Clarke, 2022). Numerous patients with CKD are at an increased risk for hyperkalemia and hyperphosphatemia, as well as the resulting chronic metabolic acidosis, bone degeneration, irregular blood pressure, and edoema. These risks can be mitigated through careful monitoring of protein, phosphorus, potassium, sodium, and calcium, thereby alleviating CKD patients’ symptoms (Naber and Purohit, 2021).

Chronic inflammation (as indicated by elevated CRP levels) is another crucial aspect of end-stage ESRD and dialysis; it contributes to a uraemic phenotype that includes cardiovascular disease, protein energy wasting, osteoporosis, and frailty (Cobo et al., 2018). Moreover, inflammatory processes contribute to low zinc (Maes, M et al., 1997), erythrocyte disorders that characterise inflammatory anaemia (Maes, M et al., 1997), and low calcium is often a sign of inflammation (Al-Hakeim et al., 2022).

### ESRD and neuropsychitric symptoms

The second major finding of this study is the significant increase in all neuropsychiatric symptom domains in ESRD patients compared to controls, as well as the significant increase in physiosomatic, fibromyalgia, and fibro-fatigue scores in patients with extreme-ESRD compared to those with late-stage ESRD. This demonstrates that ESRD patients experience a variety of mental symptoms, such as depression and anxiety, cognitive impairments, insomnia, and physiosomatic symptoms, such as fatigue, fibromyalgia, and autonomic symptoms. In CKD and ESRD, an increased prevalence of depressive symptoms and fatigue has previously been reported (Afshar et al., 2012; Karakan et al., 2011; Yoong et al., 2017).

Patients with ESRD and those undergoing long-term dialysis commonly experience fatigue and exhaustion as their most prominent symptoms (Artom et al., 2014; Chan et al., 2016; Flythe et al., 2018; Horigan et al., 2013; Jhamb et al., 2009; Manns et al., 2014). In addition, more than fifty percent of patients undergoing long-term hemodialysis suffer from fatigue symptoms (Horigan, 2012; Zyga et al., 2015), including lack of physical energy, physical and mental fatigue, cognitive impairments, and reduced day-to-day activities (Heiwe et al., 2003; Horigan et al., 2013; Lee et al., 2007).

Depression is frequently associated with fatigue in dialysis patients (Bossola et al., 2009; Farragher et al., 2017; Lee et al., 2007), whereas insomnia is linked to increased fatigue (Jhamb et al., 2009) and exhaustion (Jacobson et al., 2019). Some studies have shown that the prevalence of depression is higher in patients with CKD/ESRD than in patients with other chronic diseases (Baumeister and Harter, 2007; Ormel et al., 2007). Compared to patients with other diseases, ESRD patients exhibit a high prevalence of both depressive and anxious symptoms (Cukor et al., 2007; Preljevic et al., 2011). More than a third of hemodialysis patients in one study had anxiety disorders (Ng et al., 2015). CKD patients frequently exhibit fibromyalgia symptoms such as chronic musculoskeletal pain, muscle weakness, cramps, and insomnia (Caravaca et al., 2016). In fact, fibromyalgia symptoms (including muscle cramps) are among the top three most frequently reported physiosomatic symptoms of end-stage renal disease (ESRD) (Afshar et al., 2012).

Importantly, we discovered that the affective and physiosomatic symptoms associated with ESRD were strongly intercorrelated to the extent that a single factor could be extracted from all domains (except perhaps insomnia). The rating scales employed in this study, namely the FibroFatigue, BDI-II, and HAMA rating scales, include both “pure” mood (e.g., feelings of guilt and sadness) and physiosomatic symptoms. Due to the high impact of somatic symptoms on these scores, it is difficult to interpret correlations between biomarkers and BDI/HAMA scores as measures of severity of mental symptoms (Al-Hakeim et al., 2022c). Therefore, it is essential to calculate the relationships between biomarkers and pure mood symptoms as opposed to pure somatic symptoms. Nevertheless, after dissecting the affective and physiosomatic dimensions, we were able to demonstrate that a single latent vector could be extracted from all domains, which we termed the physio-affective phenome.

This suggests that shared pathways may explain the affective and physiosomatic symptoms of ESRD. In addition, we were able to demonstrate that affective and physiosomatic symptoms belong to the same shared core in a variety of other medical conditions, including acute COVID-19 infection, Long COVID, schizophrenia, type 2 diabetes mellitus, and unstable angina (Al-Hakeim et al., 2022a; Al-Hakeim et al., 2022b; Al-Hakeim et al., 2022c; Al-Hakeim et al., 2021). Consequently, mental (psychiatric) and somatic (physiosomatic) symptoms are intrinsically linked and driven by various ESRD-related pathways, as discussed below.

### Biomarkers of affective and physiosomatic symptoms of ESRD

The third significant finding of this study is that PLS analysis demonstrated that the physio-affective phenome of ESRD (including affective, cognitive and physiosomatic symptoms) was best predicted by the cumulative effects of aberrations in the erythron, kidney function tests, copper, zinc, and CRP, wherein the effects of kidney function decline were partially mediated by disorders in the erythron and increased copper, whereas the effects of dialysis were completely mediated by zinc and CRP. This suggests that renal dysfunctions may influence the phenotype of ESRD via mechanisms other than those involving RBC- and copper-mediated processes.

Increased inflammatory responses (including elevated CRP levels) and decreased zinc (inflammatory and antioxidant biomarker) are hallmarks of major depression (Maes, 1995; Maes et al., 1994; Siwek et al., 2010), whereas decreased calcium and increased copper are associated with affective symptoms in other conditions, including Long COVID, type 2 diabetes mellitus, and unstable angina (Al-Hakeim et al., 2022d; Maes et al., 2022; Mousa et al., 2022). As stated in the introduction, erythron disorders are also associated with major depressive disorder and chronic fatigue syndrome (see Introduction). Previously, we discussed how immune-inflammatory pathways, oxidative stress, and decreased antioxidant defences (including decreased zinc and calcium) may contribute to the pathophysiology of major depression, chronic fatigue syndrome, and physiosomatic symptoms, including those caused by other medical conditions (Al-Hakeim et al., 2022d; Leonard and Maes, 2012; Morris and Maes, 2013, 2014; Mousa et al., 2022; Maes et al., 2006). Increased copper levels and subsequent copper toxicity may cause not only physiosomatic symptoms such as nausea, gastro-intestinal, and pain symptoms, but also cognitive deficits, brain fog, and affective symptoms (Tokarz, 2021). In addition, copper modulates neurotransmission and synaptic configuration (Opazo et al., 2014), and excessive copper deposition within neurons can disrupt cortical function and exacerbate dementia symptoms (Scheiber et al., 2017).

Our results indicate that erythron abnormalities may contribute to the pathophysiology of the physio-affective phenome associated with ESRD. In ESRD, the effects of erythron on physiosomatic symptoms are likely due to an amplification of the nitro-oxidative and inflammatory pathways. Thus, increased oxidative stress and inflammation may suppress the antioxidant defences in RBCs and alter the morphometric features and functions of haemoglobin, whereas antioxidant administration may mitigate these changes (Revin et al., 2019). In addition, hemoglobin serves as an antioxidant and, therefore, the sharply decreased haemoglobin levels associated with ESRD attenuate the antioxidant defences (Jóźwik et al., 1997). As such, the lowered antioxidant capacity of RBCs in ESRD will result in increased oxidative stress (Groen et al., 2016), which is a major pathway in chronic fatigue syndrome and affective symptoms (Maes et al., 2012; Morris and Maes, 2013). Thus, disturbances in the RBC antioxidant capacity, along with high peripheral oxidative stress, may increase the vulnerability to oxidative damage and contribute to ischaemic tissue damage (Groen et al., 2016) and, consequently, to chronic fatigue and affective symptoms. Moreover, erythron disorders caused by inflammation may activate the hypoxia-inducible factor (HIF) pathway, resulting in hypoxic damage (Deng et al., 2016), as has been repeatedly reported in fatigue, depression, and anxiety (Zhao et al., 2017).

Psychologists and psychiatrists frequently attribute affective and “psycho”somatic symptoms in ESRD to negative emotions, perceived threats of long-term dialysis treatment, and the perception of depression (Lee et al., 2007). These symptoms are also attributed to fear of death, the inability to live a normal life and engage in typical hobbies, sex, creative functions, travel, and other activities, the high cost of therapy, complications of ESRD, side effects of medications used to treat CKD, and comorbidities associated with chronic kidney disease (Heiwe et al., 2003; Lee et al., 2007). Nevertheless, our results show that affective symptoms, fatigue and fibromyalgia are part of the physio-affective phenome which is largely predicted by a multitude of pathways associated with CKD and ESRD.

## Limitations of the study

In light of the findings, it is important to recognise the research’s limitations. Due to our use of a case-control design, it is impossible to draw firm conclusions about cause and effect. One could argue that the sample size is quite small. Nonetheless, an a priori calculation of the sample size revealed that a sample size of 98 is required to achieve a power of 0.80. In addition, the regression of the phenome features on the biomarkers revealed that, given the sample size, alpha=0.05, and 5-6 predictors, the actual obtained power in our study was 0.998 (for insomnia) and 1.00 (for all other regressions).

## Conclusions

ESRD is characterised by elevated scores of depressive, anxious, cognitive, and physiosomatic symptoms. One latent vector could be extracted from these diverse symptom domains, which are therefore manifestations of the physio-affective phenome. RBC abnormalities, kidney function tests, C-reactive protein, zinc, and copper together accounted for 85.0% of the variance in the physio-affective phenome. Moreover, the effects of declining kidney function on the phenome were partially mediated by RBC aberrations and elevated copper, whereas the effects of dialysis frequency were entirely mediated by decreased zinc and elevated CRP. Affective (depression and anxiety), cognitive, and physiosomatic symptoms (including chronic fatigue and fibromyalgia) are interconnected manifestations of the physio-affective phenome, which is driven by kidney dysfunctions and associated deficits in the erythron, inflammation, lowered antioxidant levels (zinc), and elevated toxicity (increased copper).

## Data Availability

All data produced in the present work are contained in the manuscript

## Acknowledgment

We thank the staff of the Dialysis Unit at Al-Hakeem General Hospital for helping in collecting samples and for Asia Clinical Laboratory in Najaf for hematological and biochemical testing.

## Author’s contributions

All the contributing authors have participated in the preparation of the manuscript.

## Data availability Statement

The dataset generated during and/or analyzed during the current study will be available from the corresponding author (MM) upon reasonable request and once the dataset has been fully exploited by the authors.

